# Estimate number of individuals infected with the 2019-novel coronavirus in South Korea due to the influx of international students from countries with virus risk: a simulation study

**DOI:** 10.1101/2020.02.15.20023234

**Authors:** Sukhyun Ryu, Sheikh Taslim Ali, Jun-sik Lim, Byung Chul Chun

**Author notes:** **Address for Correspondence:** Professor. Byung Chul Chun, Department of Preventive Medicine, Korea University College of Medicine, 73 Inchon-ro, Seongbukgu, Seoul, 02841, Republic of Korea.

## Abstract

**Background:** In March 2020, overall, 37,000 international students from the country at risk of the 2019-novel coronavirus (COVID-19) infection will arrive in Seoul, South Korea. Individuals from the country at risk of COVID-19 infection have been included in a home-quarantine program, but the efficacy of the program is uncertain.

**Methods:** To estimate the possible number of infected individuals within the large influx of international students, we used a deterministic compartmental model for epidemic and perform a simulation-based search of different rates of compliance with home-quarantine.

**Results:** Under the home-quarantine program, the total number of the infected individuals would reach 24–53 from March 17–March 20, 50–86 from March 18– March 16, and 234– 343 from March 4– March 23 with the arrival of 0.1%, 0.2%, and 1% of pre-infectious individuals, in Seoul, South Korea, respectively. Our findings indicated when incoming international students showed strict compliance with quarantine, epidemics were less likely to occur in Seoul, South Korea.

**Conclusion:** To mitigate possible epidemics, additional efforts to improve the compliance of home-quarantine are warranted along with other containment policies.

## BACKGROUND

Three major respiratory virus-related events have been observed in South Korea in the 21^st^ century: severe acute respiratory virus (SARS), Middle East respiratory syndrome, and the 2019-novel coronavirus (COVID-19) infection, all of which are caused by members of the coronavirus family. The first individual with COVID-19 infection in South Korea was identified on January 20, 2020 and, the number of laboratory-confirmed cases increased between then and February 12, 2020 [1]. To reduce the number of individuals entering South Korea who may have been exposed to COVID-19 in Wuhan, China, an international travel ban from Hubei Province, China to South Korea was implemented on February 3, 2020 (Figure 1) [2]. Furthermore, to identify individuals who may have been exposed to COVID-19, the South Korean public health authority implemented a quarantine program. Any persons who have travelled from a country with COVID-19 infection risk within the previous 14 days or have been in contact with laboratory-confirmed COVID-19 infection within the previous 14 days is defined as an individual for quarantine [3]. Quarantined individuals are asked to comply with home-quarantine and are monitored by local public health workers twice a day for 14 days after contact with individuals with infection [3].

**Figure 1.**
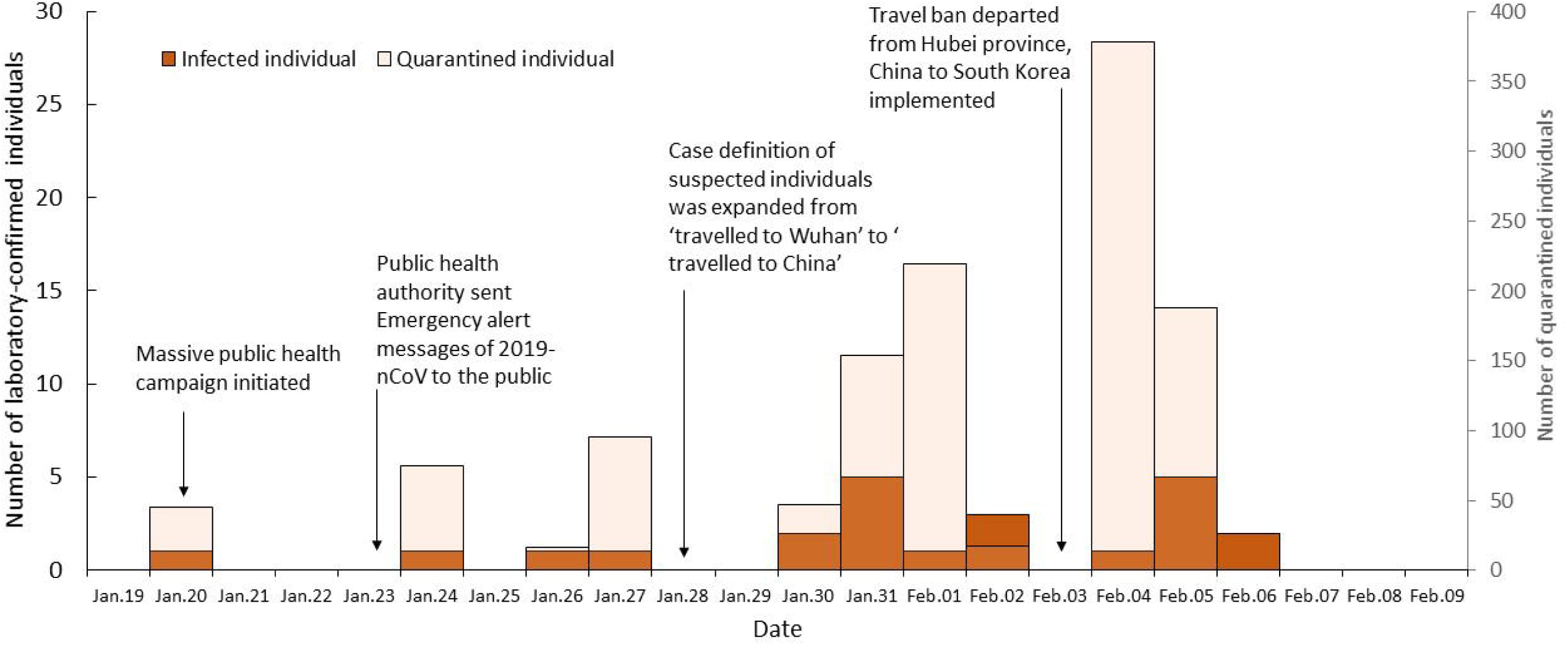
Timeline of the number of laboratory-confirmed cases and the number of quarantined individuals with 2019-novel coronavirus infection in South Korea

On February 14, 2020, the South Korean public health authority identified an individual with COVID-19 infection; the patient had been contacted by another individual who was suspected of avoiding the quarantine program during his period of home-quarantine [4]. According to previous literature, the effectiveness of quarantine varies widely depending on individuals’ daily motility patterns [5]; Despite this compliance with home-quarantine in the present instance is still in question.

It is important to note that 37,000 students from China, where major cities are likely experiencing localized outbreaks [6], will enter Seoul, South Korea, on March 1, 2020 at the start of the spring semester. This large number of incoming youths from the country with COVID-19 infection risk may increase the risk of local transmission in South Korea.

In this study, we aimed to estimate the number of infected individuals in Seoul, South Korea, based on compliance with home-quarantine among these incoming international students.

## METHODS

To simulate possible epidemics, we used the deterministic compartmental model of the susceptible-exposed-infectious-removed type (see the Supplementary Appendix). We assumed that the population mixed homogeneously, and that no COVID-19 transmission had occurred within the community in Seoul, South Korea. We assumed that either 0.1%, 0.2%, or 1% of the incoming international students were in the pre-infectious period of COVID-19 infection, based on previous literature reporting that 0.2% of individuals with contactees of SARS infection were asymptomatic [7]. We also assumed that the international students would arrive in Seoul, South Korea in the 15 days before and after March 1, 2020, and that no individuals were isolated during entry screening upon arrival. Furthermore, we assumed that all quarantined individuals were confined at home or to the university dormitory as per the current South Korean quarantine program for COVID-19 infection implemented by the local public health authority. The baseline scenarios were based on the currently identified number of infected persons from China in South Korea, which was 12 on February 6, 2020, with the assumption of 90% compliance with home-quarantine during the pre-infectious period. Scenarios with different quarantine compliance rates (70%, 80%, or 90%) among these international students were also modeled. We considered a time horizon of 180 days for the number of individuals infected and quarantined since January 20, 2020, when the first COVID-19 case was identified in South Korea. The parameter values of our model, obtained from previous studies, are shown in the Supplement Material.

## RESULTS

We estimated that the total number of infected individuals would reach 24–53 from March 17–March 20, 50–86 from March 18–March 16, and 234–343 from March 4–March 23 with the arrival of 0.1%, 0.2%, and 1% of pre-infectious individuals, in Seoul, South Korea, respectively (Figure 2).

**Figure 2.**
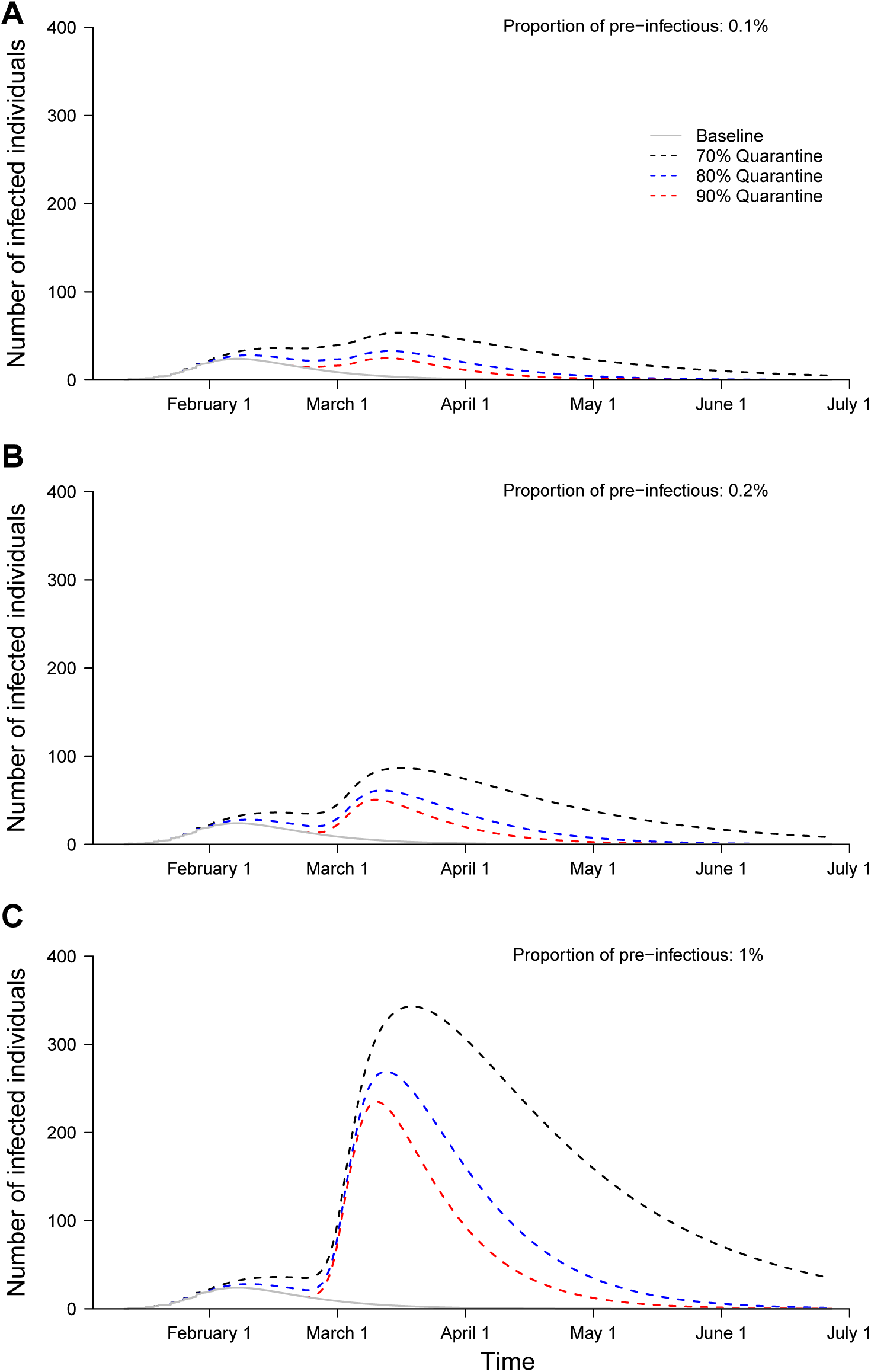
Estimated daily number of individuals with infection in Seoul, South Korea under different scenarios regarding the proportion of pre-infectious individuals: 0.1% (a), 0.2% (b), and 1% (c), based on different compliance rates with home-quarantine (gray: baseline, black: 70%, blue: 80%, red: 60%).

We also estimated that the number of individuals isolated from the South Korean quarantine program would peak at 24–47 from March 17–March 25, 48–77 from March 16– March 28, and 225–305 from March 14–March 25 with the arrival of 0.1%, 0.2%, and 1% of pre-infectious individuals in Seoul, South Korea, respectively (Figure 3). The number of infected and isolated individuals would increase with higher proportions of subclinical COVID-19 cases. However, the number of infected and isolated individuals was smaller due to the high compliance of the quarantine program.

**Figure 3.**
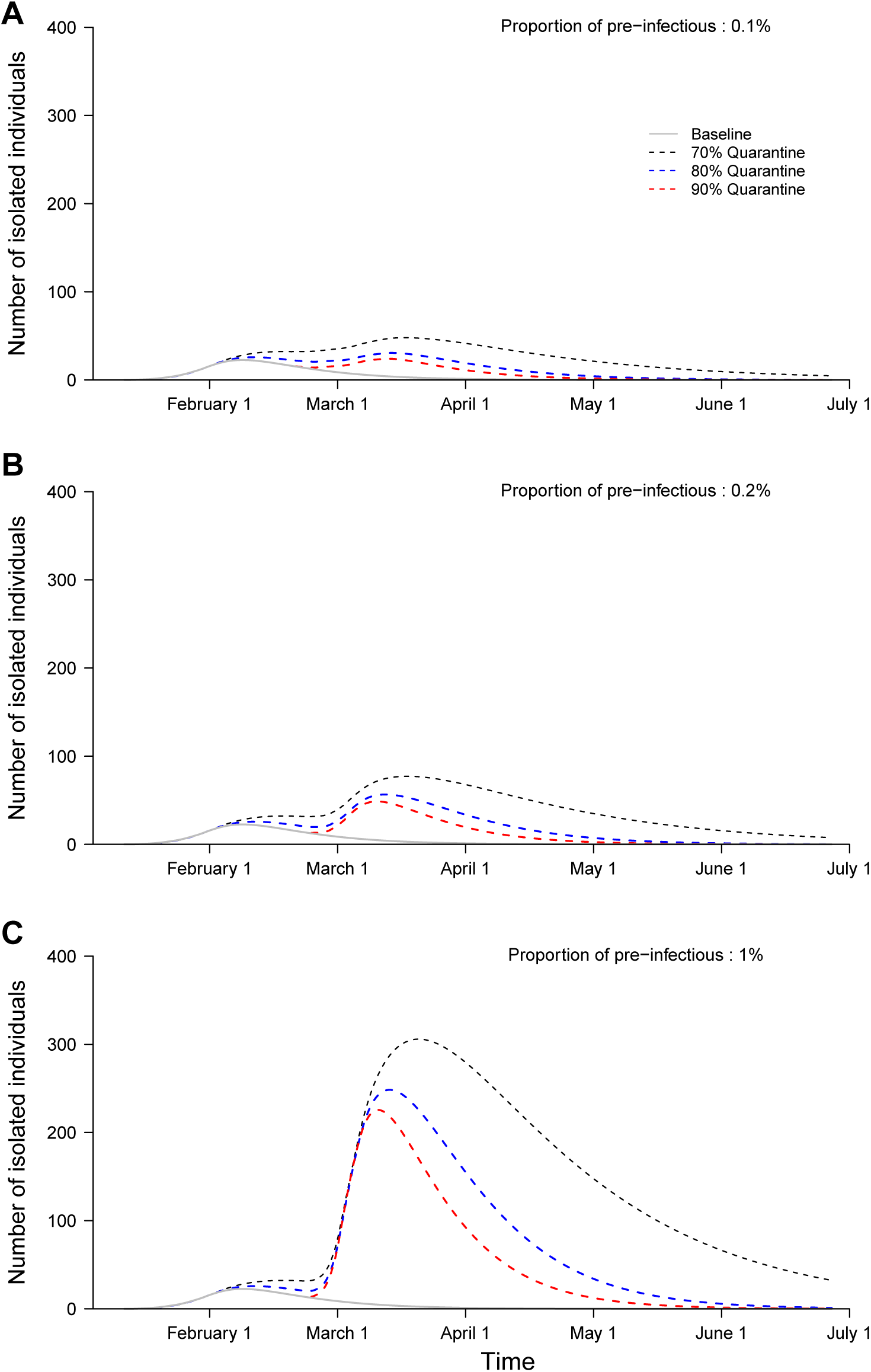
Estimated daily number of isolated individuals in Seoul, South Korea, under different scenarios regarding the proportion of pre-infectious individuals: 0.1% (a), 0.2% (b) or 1% (c) based on different compliance rates with home-quarantine (gray: baseline, black: 70%, blue: 80%, red: 60%).

## DISCUSSION

When no effective vaccine or treatment is available for infectious disease, the quarantine of individuals suspected of having the infection, including those exposed to infection from epidemic countries, has been used as a mitigation strategy by public health authorities [8, 9].

The number of laboratory-confirmed individuals with COVID-19 infection is increasing in China and other Asian countries. In South Korea, the likelihood of local transmission is increasing because travelers are arriving from COVID-19-affected countries. The quarantine of individuals who may have been exposed to COVID-19 is an efficient public health strategy, to reducing transmission while using limited public health resources, because the presence of individuals with unidentified infection is highly likely among individuals exposed to the infectious diseases [9]. Therefore, the number of individuals with infection can be estimated based on compliance with home-quarantine to provide relevant evidence for public health authorities and to improve international students’ compliance with the quarantine program in advance.

In South Korea, individuals who had contacted a person with infection were asked to comply with home-quarantine and were monitored by local public health workers twice a day for 14 days post-contact [3]. Individuals who were not included in the quarantine program but had experienced any possible contact were encouraged to notify public health authorities and submit to quarantine. All daily necessities were provided to all quarantined individuals by the public health authorities to avoid possible contact with any susceptible population, as indicated by the South Korean law. Therefore, the current quarantine program in South Korea is very broad and includes a large number of people. However, to relieve the pressure on public health resources, the quarantine program for incoming international students will be monitored by the education authority [10]. This may affect the efficacy of quarantine and increase the number of infected and isolated individuals.

Our findings indicate that most of the infected individuals isolated from the home-quarantine program; Therefore, epidemics by incoming international students are unlikely to occur in Seoul, South Korea; However, the number of infected and isolated individuals could increase by mid or late March. Furthermore, the quarantine program may consume a large number of public health resources because it involves monitoring quarantined individuals and isolating infected individuals.

The present study had several limitations. Firstly, some parameters including the latent period and rate of infection among those in contact with a person with infection were obtained from the modelling studies of COVID-19 [6, 11, 12], and consequently may revise the results. Secondly, we used a deterministic model, and can’t evaluate the uncertainty of these estimates, which is an inherent feature and missed under current analysis. However, allowing a search of different plausible values of these parameters through this model simulation approach, ensures the reliable parameter estimates and able to mimic the future dynamics of the number of infected individuals, which is much smaller than the total population [13]. Thirdly, we did not consider transmission that occurred before symptom onset and did not account for subclinical infection.

## CONCLUSIONS

As public health resources are limited, quarantine of individuals who may have been exposed to infectious disease is crucial for preventing local transmission [14]. Therefore, strict home-quarantine of individuals from countries at risk for COVID-19 infection is important to reduce the number of infected individuals and to prevent possible epidemics in the community.

## Data Availability

Not applicable

## Declarations

### Ethics approval and consent to participate

Not applicable

### Consent for publication

Not applicable

### Author Contributions (Use CRediT terms)

Conceptualization: SR; Methodology: SR, and STA; Formal analysis: SR, and JL; Data curation: SR; Validation: SR, STA, and JL; Writing - original draft preparation: SR, and BCC; Writing - review and editing: SR, JL, and BCC; Approval of final manuscript: all authors.

## ACKNOWLEDGMENTS

SR reports a past research grant from Basic Science Research Program through the National Research Foundation of Korea by the Ministry of Education (grant number NRF-2018R1A6A3A03012236) and Mogam Science Scholarship Foundation.

## Author’s information

Dr. Ryu is an assistant professor of preventive medicine at Konyang University, Daejeon, South Korea. His research interests include infectious disease epidemiology, with a focus on influenza and public health interventions.

## ORCID

Sukhyun Ryu https://orcid.org/0000-0002-8915-8167

Sheikh Taslim Ali https://orcid.org/0000-0002-8631-9076

Jun-sik Lim https://orcid.org/0000-0003-4645-2347

Byung Chul Chun https://orcid.org/0000-0001-6576-8916

